# Which African Countries are at Risk of Missing SDG 3.2? Bayesian Mapping of Under-Five Mortality Using UNICEF 2024 Data

**DOI:** 10.64898/2026.07.04.26357223

**Authors:** Oladimeji Damilare Matthew, Mustapha Kamilu Adisa, Ekop Eloho Eno

## Abstract

**Background:** Despite considerable reductions in under-five mortality during the Millennium Development Goal era, progress towards Sustainable Development Goal (SDG) 3.2 remains uneven across Africa. Identifying countries at greatest risk of missing the target is essential for prioritizing interventions and resource allocation.

**Methods:** A Bayesian spatial forecasting ecological study was conducted using 2024 country-level data from 49 African countries obtained from UNICEF. Spatial dependence was assessed using Global Moran’s I and Local Indicators of Spatial Association. Bayesian structured additive regression models with Gaussian, Gamma, and Exponential likelihoods were fitted using Integrated Nested Laplace Approximation (INLA) and compared using the Deviance Information Criterion (DIC), Watanabe-Akaike Information Criterion (WAIC), and conditional predictive ordinates. Posterior exceedance probabilities were estimated, an SDG Failure Index (SFI) and a Priority Intervention Index (PII) were developed, and Bayesian posterior predictive simulations were performed to estimate country-specific probabilities of attaining SDG 3.2 by 2030.

**Results:** Significant spatial clustering of under-five mortality was observed with (Moran’s I = 0.355, *p* < 0.001), and hotspots in Benin, Cameroon, and Nigeria. The Gamma model provided the best fit (DIC = 114.92; WAIC = 111.71). Diarrhoea was the only significant predictor (posterior mean=0.030; 95% credible interval: 0.004-0.056). Twenty-three countries (46.9%) were classified as high risk, whereas only five (10.2%) had achieved SDG 3.2. West Africa recorded the highest mean mortality (7.05%) and North Africa the lowest (1.64%). Bayesian projections indicated that only five countries were likely to achieve SDG 3.2 by 2030, while 41 (83.7%) were unlikely to do so.

**Conclusion:** Considerable geographical inequalities in under-five mortality persist across Africa, and most countries remain off-track for achieving SDG 3.2 by 2030. The integration of exceedance probability mapping, the SDG Failure Index, the Priority Intervention Index, and Bayesian probability forecasting provides a practical framework for monitoring progress and prioritizing countries requiring accelerated action towards achieving SDG 3.2.

## Introduction

Under-five mortality is widely regarded as a sensitive indicator of population health because it captures the combined influence of maternal well-being, household living conditions, nutrition, environmental exposures, and access to quality healthcare services on child survival [1]. Over the last three decades, significant progress has been recorded in reducing child mortality worldwide. According to 2024 United Nations Inter-agency Group for Child Mortality Estimation (UN IGME), the global under-five mortality rate declined from 93 deaths per 1,000 live births in 1990 to 37 deaths per 1,000 live births in 2023[2]. Despite this progress, the burden of preventable child deaths remains disproportionately concentrated in low and middle-income countries, with sub-Saharan Africa continuing to experience the highest mortality rates and accounting for a substantial share of global under-five deaths burden[2]. Recent evidence also suggests that the pace of improvement has slowed in several settings, raising concerns about the likelihood of achieving global child-survival targets within the stipulated timeframe [2]. The reduction of preventable deaths among children under five forms an important component of the Sustainable Development Goals (SDGs). Specifically, SDG 3.2 aims to reduce under-five mortality to fewer than 25 deaths per 1,000 live births by the year 2030 [3]. While a number of African countries have made notable progress toward this target, achievements have not been the same across the continent. Noticeable differences in mortality levels still persist between countries and regions, reflecting disparities in socioeconomic development, healthcare infrastructure, maternal and child health services, and broader social determinants of health [4].

Previous investigations have overtime claimed that under-five mortality has a link with factors such as poor maternal health, inadequate immunization, malnutrition, infectious diseases, poverty, and limited access to healthcare services [1,4,5]. Other factors like stunting, undernutrition, diarrhoeal diseases, pneumonia, and complications associated with maternal ill-health continue to contribute handsomely to child deaths across many African countries [5,6]. Beyond differences in mortality levels, research increasingly demonstrates that child mortality is not distributed uniformly across geographical areas. Countries that share common socioeconomic, environmental, and health-system characteristics often exhibit similar mortality patterns, resulting in spatial clustering of child survival outcomes [7,8]. Coming to terms with these geographic patterns is important because national averages may conceal areas where child mortality remains persistently high. Spatial epidemiological methods provide an opportunity to identify such inequalities and generate evidence that can guide geographically targeted interventions. In this regard, Bayesian spatial models have gained considerable attention because they explicitly account for spatial dependence, produce stable risk estimates, and provide comprehensive measures of uncertainty [9]. The Integrated Nested Laplace Approximation (INLA) framework has further enhanced the practical application of Bayesian modelling by enabling efficient estimation of complex spatial models without the computational burden associated with traditional simulation-based approaches [10]. Although the literature on under-five mortality in Africa has expanded considerably, several knowledge gaps remain. Much of the existing evidence is derived from single country studies or analyses conducted at subnational levels, making it difficult to obtain a comprehensive understanding of mortality inequalities across the African continent as a whole [5,7]. In addition, previous studies have predominantly focused on describing mortality patterns or identifying associated risk factors[8]. Comparatively less attention has been devoted to evaluating the probability that countries remain above internationally accepted mortality thresholds. From a policy perspective, such information is critical because countries with similar mortality rates may differ substantially in their likelihood of meeting global targets. Exceedance posterior probability mapping provides a useful framework for addressing this issue by quantifying the probability that mortality exceeds a specified benchmark and highlighting locations where the risk of failing to achieve development goals is greatest [9]. Against this background, this present study investigates the spatial distribution of under-five mortality across Africa using UNICEF 2024 data within a Bayesian spatial modelling framework. The study employs Integrated Nested Laplace Approximation to estimate spatially smoothed mortality risks, examine geographical clustering, assess factors associated with mortality variations, and determine the probability that individual countries exceed the SDG 3.2 target of 25 deaths per 1,000 live births.

Although substantial efforts have been made to monitor progress toward SDG target 3.2, most existing assessments rely primarily on observed mortality levels or deterministic projections based on historical trends. Such approaches provide limited information regarding the uncertainty surrounding future attainment, the magnitude of expected shortfalls, and the relative urgency of interventions required across countries. Recent studies have increasingly explored probabilistic and Bayesian approaches for forecasting child survival outcomes and assessing prospects for achieving SDG targets in Africa[8]. However, available frameworks seldom integrate uncertainty quantification, distance from the SDG target, and the pace of mortality reduction within a single analytical framework.

The annual rate of reduction (ARR) is widely used as a summary measure for evaluating progress in child mortality reduction because it reflects the speed at which countries are moving toward internationally agreed targets. Nevertheless, countries exhibiting similar mortality burdens may differ substantially in their rates of decline, implying different prospects for attaining SDG target 3.2 by 2030. To address these challenges, the present study combines ARR with Bayesian posterior predictive simulation and composite prioritization metrics to provide a more comprehensive assessment of child survival prospects in Africa.

Specifically, the study estimated country-specific probabilities of attaining the SDG target by 2030 through Bayesian posterior simulation, quantified expected shortfalls using an SDG Failure Index (SFI), and developed a Priority Intervention Index (PII) that jointly considers the magnitude of expected non-attainment and the prevailing pace of mortality decline. These complementary measures extend conventional SDG monitoring beyond descriptive comparisons and offer an evidence-based framework for identifying countries where accelerated investments and policy actions may yield the greatest gains in child survival.

## Methods

### Study Design

This study employed a Bayesian spatial ecological forecasting design to investigate geographical variation in under-five mortality, quantify uncertainty of attaining SDG target 3.2 and evaluate prospects for achieving SDG 3.2 across 49 African countries.

### Data Source

Data for this study were obtained from the United Nations Children’s Fund(UNICEF) 2024 database. UNICEF provides internationally harmonized indicators on Child health, nutrition, maternal health, immunization coverage and healthcare services for countries worldwide. The database is widely used for monitoring progress towards child survival targets and the SDG Goals. Spatial boundary data for African countries were obtained from publicly available geographic information system(GIS) shapefiles.

### Study variables

The study consisted of one outcome variable and a set of explanatory variables selected. The dependent variable was Under-five mortality rate(U5MR) which was defined as the probability of a child dying before reaching five years of age[2]. U5MR serves as a key indicator of child survival and is used globally to monitor progress toward SDG 3.2[2]. The independent variables were selected based on theoretical relevance and empirical evidence linking them to child mortality outcome. The selection of these variables was guided by child survival framework proposed by Mosley and Chen (1984) which emphasizes the influence of maternal factors, nutritional status, disease burden, healthcare financing condition on child mortality.

### Ethics statement

This study involved the analysis of secondary datasets from the UNICEF dataset 2024. The datasets obtained for analysis were anonymised and all unique identifiers were removed before the release of the data for public use.

### Model Specification

The structured additive predictor is defined as

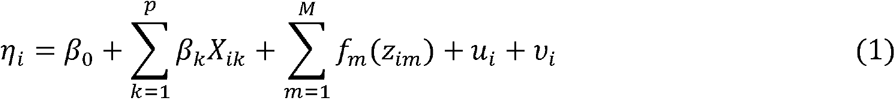

Where;*β*_0_ = *intercept,β*_*k*_ = *fixed effect coefficient; X*_*ik*_ = *linear covariates; f*_*m*_(·) = *nonlinear smooth effects; u*_*i*_ = *structred spatial effect; v*_*i*_ = *unstructured random effect*.

For this study;

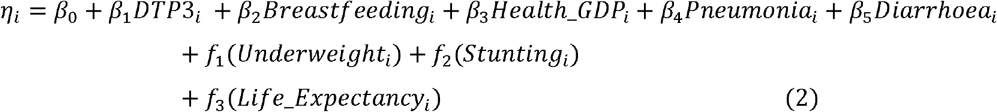

Spatial Dependence structure

Let W = (*w*_*ij*_) be the adjacency matrix:

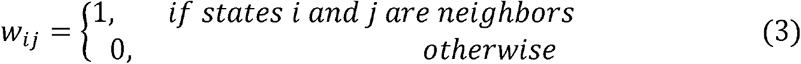

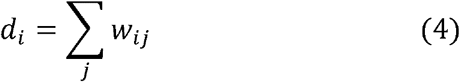

Conditional distribution

The intrinsic CAR model specifies:

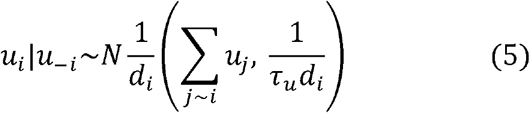

Mean=average of neighboring states

Variance decreases as number of neighbors increases.

Joint Distribution

The joint distribution of spatial effects is:

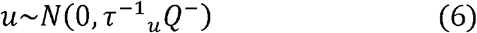

where

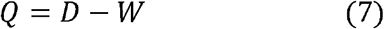

D diagonal matrix with entries *d*_*i*_

Q=precision matrix

Because Q is singular, this defines an intrinsic CAR(ICAR) model.

The CAR prior enforces *u*_*i*_ ≈ *u*_*j*_ for neighboring regions

Thus, it captures spatial clustering, similar U5MR behaviour in nearby countries and unobserved regional influences

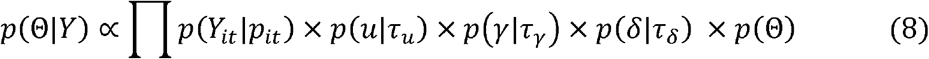

this model is estimated using Integrated Nested Laplace Approximation (INLA), which provide accurate and computationally efficient Bayesian Inference for latent Gaussian or any appropriate models.

### Exceedance Probability Analysis

The SDG 3.2 threshold correspond to c=2.5 representing 25 deaths per 1000 livebirths. Posterior exceedance probabilities were estimated as

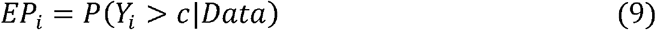

Countries *EP*_*i*_ > 0.8 were considered highly likely to remain above the SDG threshold.

### SDG Gap Analysis

The SDG gap for country i was computed as

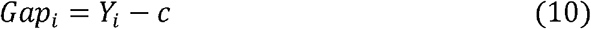

Where *Y*_*i*_ denotes the observed under-five mortality rate

Positive values imply mortality burden above the SDG target while negative values indicate target attainment.

### SDG Failure Index

To simultaneously quantify magnitude of deviation from SDG target 3.2 and the uncertainty associated with target attainment, an SDG Failure Index (SFI) was developed in this study.

For country i, the SDG gap was defined in eq(10).

The SDG Failure Index was computed as

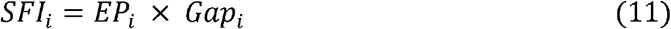

Where

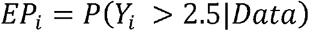

Denotes the posterior exceedance probability obtained from the Bayesian spatial model. Large values of the index indicate countries with simultaneously large mortality deficits and high posterior probabilities.

### Priority Intervention Index

To identify countries require accelerated interventions, a priority Intervention Index(PII) was proposed by combining the SDG failure Index with recent progress in mortality reduction. The annual rate of reduction (ARR) reported by UNICEF between 2015 and 2024 was incorporated as

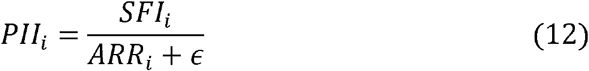

Where *ϵ* = 0.1 and was introduced to avoid numerical instability associated with countries exhibiting very small or negative reduction rates. Countries were subsequently classified into three categories see table 1.

**Table 1:**
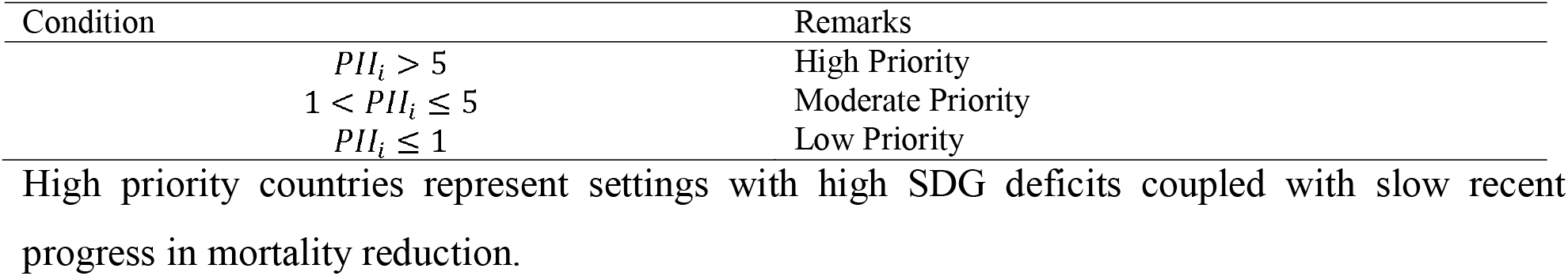
Priority Intervention Cut-off.

**Table 2:** Posterior probability Cut-off of achieving SDG 3.2.

| Condition | Remarks |
| --- | --- |
| $P_i \geq 0.80$ | Likely Achieve |
| $0.5 \leq P_i < 0.80$ | Possible |
| $P_i < 0.5$ | Unlikely |

### Bayesian Probability of Achieving SDG 3.2 by 2030

Posterior predictive simulation was used to estimate the probability of attaining SDG target 3.2 by 2030. For each country, posterior samples were generated from

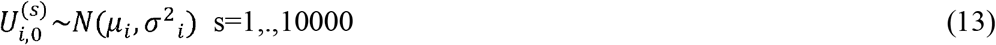

Where *μ*_*i*_ = *posterior mean and*

*σ*_*i*_ = *posterior standard deviation*

Obtained from the Bayesian Gamma INLA model. Assuming continuation of recent mortality reduction trends, projected mortality levels in 2030 were estimated as

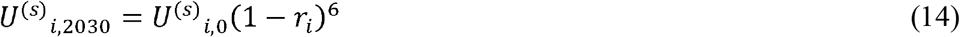

Where 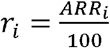 represent the annual rate of reduction and six years correspond to the interval between 2024 and 2030. The posterior probability of achieving SDG 3.2 was then calculated as

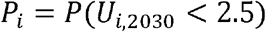

Which was estimated by

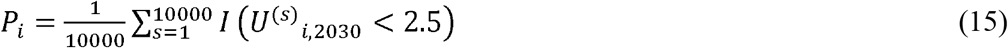

Where *I*(·) is an indicator function taking value one when the simulated mortality level satisfies the SDG target and zero otherwise. Countries were classified as

### Continental and Regional Inequality Assessment

Inequality in under-five mortality across African countries was quantified using the Gini Coefficient

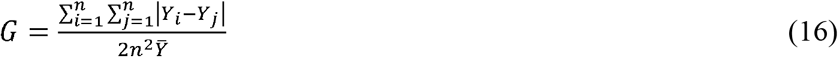

Where *Y*_*i*_ denotes the under-five mortality rate in country i;, 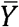 represents the continental mean under-five mortality rate; n is the number of countries included in the analysis. The Gini coefficient ranges from 0 ≤ *G* ≤ 1 with G=0 indicating perfect equality; G=1 indicating complete inequality.

## Results

### Descriptive Characteristics

The results in Table 3 indicate descriptive statistics of the variables used in this study. Overall average of Africa prevalence of U5MR is 5.3% which implies 53 per 1000 livebirth. Nigeria accounted for highest of U5MR in the region with almost 116 per 1000 livebirth follow by Niger and Chad with 111 and 97 per 1000 livebirth respectively. Libya and Cape Verde have the lowest burden in region with 10 and 11 per 1000 livebirth respectively. The average DTP3 uptake is 80.4%, Exclusive breastfeeding is 55.3%, prevalence of underweight is 14.4%, average life expectancy in the region is 67 years and average percentage of under five children out total population in the region is 13.4%

**Table3:** Summary Statistics.

| Variable | Mean | SD | Median | Minimum | Maximum |
| --- | --- | --- | --- | --- | --- |
| Under-Five Mortality Rate | 5.29 | 2.74 | 4.85 | 1 | 11.6 |
| DTP3 (%) | 80.4 | 13.6 | 83.5 | 39 | 98 |
| Exclusive Breastfeeding (%) | 55.29 | 18.37 | 56 | 17 | 93 |
| Underweight Prevalence (%) | 14.37 | 3.74 | 15 | 7 | 23 |
| Stunting Prevalence (%) | 24.85 | 11.71 | 23.5 | 5 | 55 |
| Life Expectancy (Years) | 67.24 | 6.22 | 67.75 | 54.8 | 79.3 |
| Population (%) | 13.44 | 3.14 | 13.97 | 4.71 | 18.56 |
| Health Expenditure (% GDP) | 7.3 | 3.37 | 7.15 | 1.6 | 15.8 |
| Maternal Mortality Ratio | 0.29 | 0.21 | 0.24 | 0.02 | 0.99 |
| Pneumonia (%) | 55.53 | 19.23 | 58.15 | 14 | 99.1 |
| Diarrhoea (%) | 38.83 | 17.93 | 35.7 | 9.9 | 93.6 |

### Correlation and Multicollinearity

Figure 1, display correlation heatmap of U5MR and selected predictors in this study. The Overall results indicate that U5MR is strongly associated with indicators of population health, maternal health, nutritional status, and demographic characteristics. Life expectancy exhibited the strong negative correlation with U5MR (r=-0.822), which implies that countries with higher life expectancy tend to experience substantially lower under-five mortality rates. Also, maternal mortality ratio shows high positive correlation with U5MR (r = 0.749), i.e countries with high maternal mortality burdens are likely to have elevated under-five mortality rates. Percentage of Under five children from total Population size was also positively correlated with U5MR (r=0.647), implying that more populous countries generally experience higher levels of child mortality. Stunting prevalence also showed a moderate positive correlation with U5MR (r = 0.480), suggesting that chronic childhood undernutrition is an important contributor to under-five mortality across the continent of Africa. Health expenditure as a percentage of gross domestic product exhibited a moderate negative correlation with U5MR (r = −0.507), indicating that increased investment in health systems may improve child survival. Underweight prevalence also showed a weak positive correlation with U5MR (r = 0.272).

**Figure 1:**
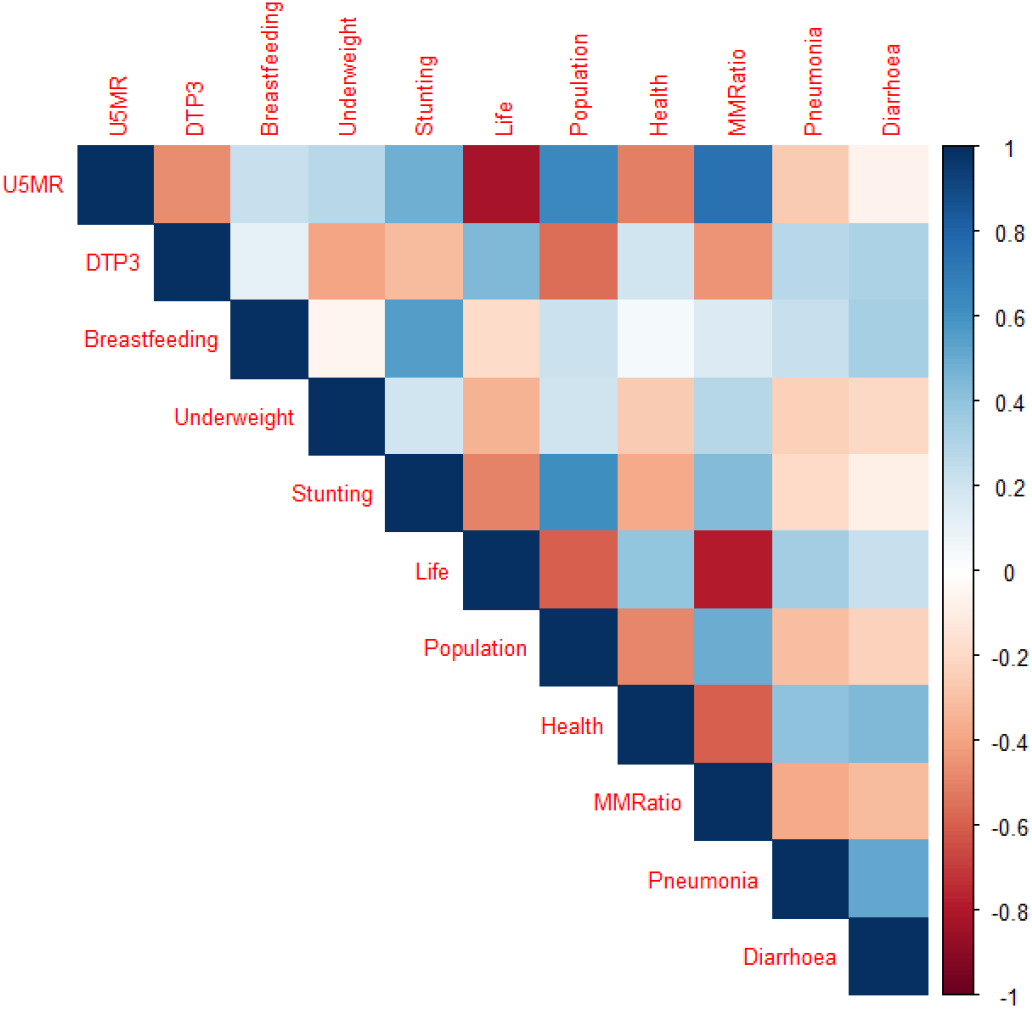
Correlation Coefficient Heatmap of USMR

### Multicollinearity

Table 4 presents the variance inflation factors (VIFs) for the explanatory variables included in the study. The estimated VIF values ranged from 1.5 to 4.1, which indicate low to moderate levels of correlation among the predictors. Maternal mortality ratio had the highest VIF value of 4.1, followed by life expectancy with (VIF = 3.6) and health expenditure with VIF of 3.13. The lowest is Pneumonia with(VIF=1.5). Since all estimated VIFs values were below the threshold of five, there was no evidence of serious multicollinearity among the explanatory variables. Consequently, all covariates were retained for subsequent Bayesian spatial modelling.

**Table 4:** Variance Inflation Factors.

| Predictor Variable | VIF |
| --- | --- |
| DTP3 Immunization | 2.27 |
| Breastfeeding | 2.2 |
| Underweight | 2.65 |
| Stunting | 2.75 |
| Life Expectancy | 3.6 |
| Population | 2.9 |
| Health Expenditure | 2.6 |
| Maternal Mortality Ratio | 4.1 |
| Pneumonia | 1.5 |
| Diarrhoea | 1.69 |

### Spatial Pattern of U5MR

Figure 2 displays the spatial distribution of the observed under-five mortality rate (U5MR) across African countries in 2024. Geographical variation in mortality burden was evident across the continent, with high concentration in mortality levels in West, Central, and parts of East Africa.

**Figure 2:**
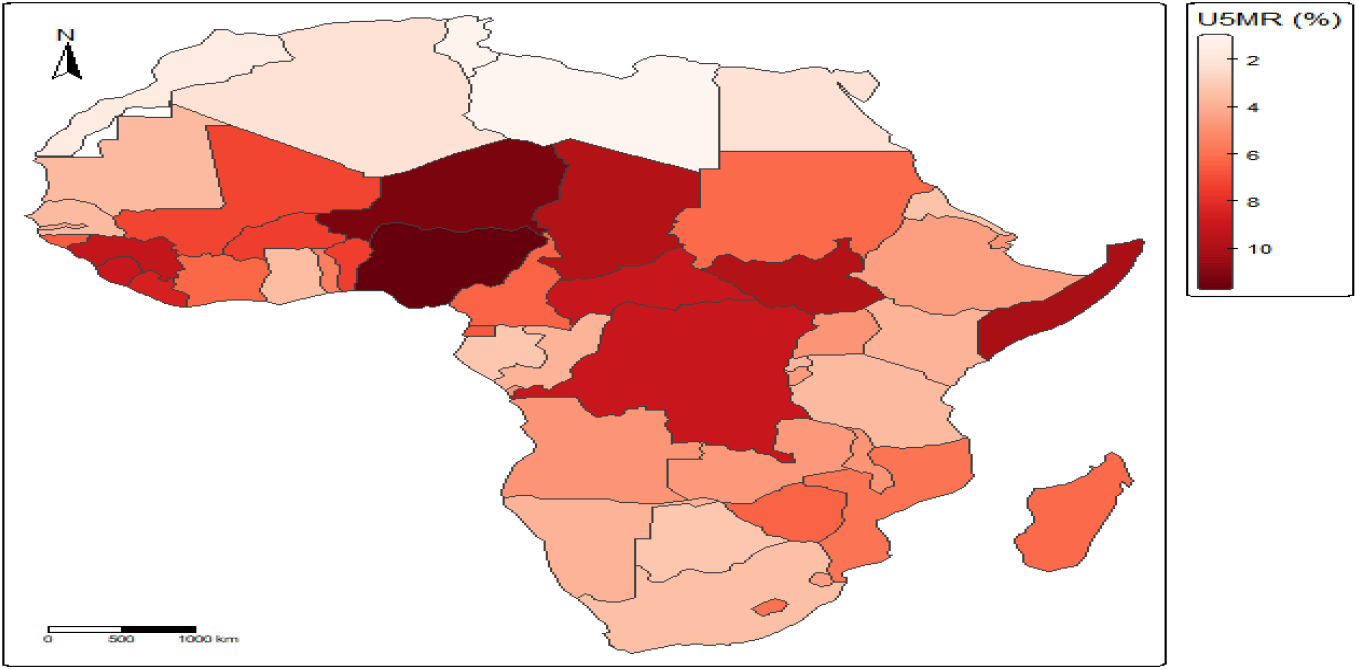
Raw prevalence map

The highest under-five mortality rates were observed in Nigeria and two of her neighouring countries Niger and Chad, and other like Somalia, South Sudan, the Democratic Republic of Congo, Guinea, Sierra Leone, Liberia, and the Central African Republic, which were depicted by the darkest shades on the map. These countries constitute a contiguous belt of elevated mortality extending from West Africa through Central Africa to the Horn of Africa. In another vein, substantially lower mortality rates were observed in North African countries, which include Libya, Tunisia, Morocco, Algeria, and Egypt, as well as some countries in Southern Africa, which were represented by lighter shades. The observed spatial pattern suggests that under-five mortality in Africa is not uniformly distributed but is characterized by distinct geographical clustering. Countries sharing common borders tended to exhibit similar mortality levels, particularly within West and Central Africa, whereas countries in North Africa demonstrated relatively favourable child survival outcomes. These preliminary observations provide visual evidence of spatial dependence and justify subsequent formal assessment of spatial autocorrelation and Bayesian spatial modelling.

### Spatial Clustering

The results of the global spatial autocorrelation analysis as presented in Table 6. Show that the estimated Moran’s I statistic was 0.355, which was higher than the expected value under spatial randomness (Expected Moran’s I = −0.021). Also, the associated variance was 0.010, and the test was highly significant at 0.05 with (p<0.001). These findings depicts the presence of significant positive spatial autocorrelation in under-five mortality across African countries, which indicates that countries with similar mortality burdens tend to be geographically clustered rather than randomly distributed across the continent.

**Table 6:** Moran I test.

| Moran's I | Expected Moran's I | Variance | p-value |
| --- | --- | --- | --- |
| 0.355 | -0.021 | 0.01 | <0.001 |

#### Hotspot analysis

To identify the locations contributing to the observed clustering pattern, Local Indicators of Spatial Association (LISA) were estimated and mapped in Figure 3. The analysis revealed three statistically significant High-High clusters comprising of Benin, Nigeria, and Cameroon. These countries exhibited high under-five mortality rates and were surrounded by neighbouring countries with similarly high mortality burdens, indicating the existence of a localized hotspot of child mortality within West and Central Africa(See table 7). In contrast, Tunisia was identified as the only significant Low-Low cluster, which indicate that the country and its neighbouring countries share relatively low levels of under-five mortality. This pattern represents a localized cold-spot of favourable child survival outcomes in North Africa.

**Table 7:** Distribution of LISA Cluster Categories for Under-Five Mortality in Africa.

| Cluster Type | Frequency | Percentage |
| --- | --- | --- |
| High-High | 3 | 6.1 |
| Low-Low | 1 | 2 |
| High-Low | 0 | 0 |
| Low-High | 0 | 0 |
| Not Significant | 45 | 91.9 |

**Figure 3:**
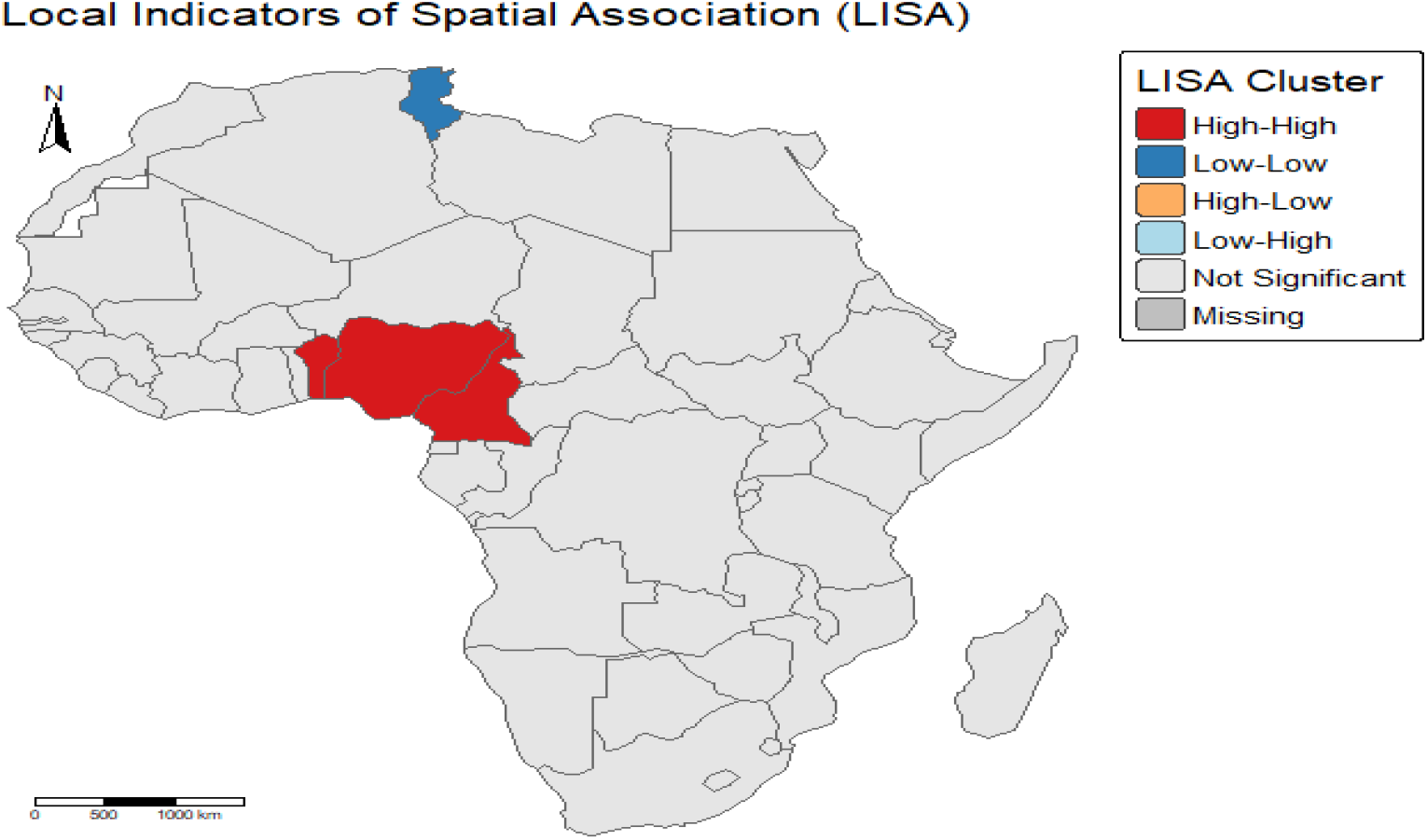
Local Indicators of Spatial Association (LISA) Cluster Map of Under-Five Mortality in Africa, 2024

### Bayesian Spatial Model

Table 8 presents the model fit statistics for the three candidate Bayesian models. The Gamma model provided the best fit to the data, yielding the lowest values of the Deviance Information Criterion (DIC =114.92) and Watanabe-Akaike Information Criterion (WAIC = 111.71). In comparison, the Gaussian model produced higher DIC (163.78) and WAIC (165.88) values, while the Exponential model demonstrated the poorest performance with substantially larger DIC (280.74) and WAIC (272.16) estimates. Although the Gaussian model had the smallest mean negative log conditional predictive ordinate (−log(CPO) = 1.75), the Gamma model was selected for subsequent analyses based on its superior overall fit as indicated by the DIC and WAIC criteria.

**Table 8:**
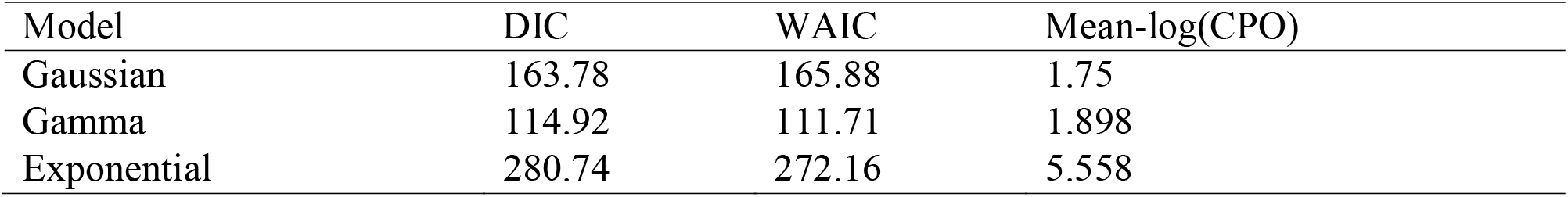
Model comparison test.

| Model | DIC | WAIC | Mean-log(CPO) |
| --- | --- | --- | --- |
| Gaussian | 163.78 | 165.88 | 1.75 |
| Gamma | 114.92 | 111.71 | 1.898 |
| Exponential | 280.74 | 272.16 | 5.558 |

#### Fixed Effect Model

The posterior estimates of the fixed effects shown in Table 9 indicate that among the covariates considered, diarrhoea prevalence was the only variable whose 95% credible interval excluded zero, suggesting evidence of an association with under-five mortality after accounting for spatial dependence and nonlinear effects. The posterior mean estimate for diarrhoea was 0.032 (95% CrI: 0.006, 0.057). The remaining fixed effects, including DTP3 immunization coverage, breastfeeding prevalence, health expenditure, and pneumonia prevalence, had credible intervals that contained zero, showing limited evidence of an association with under-five mortality.

**Table 9:** Fixed effect parameter Estimates.

| Covariate | Mean (SD) | 95% Credible Interval |
| --- | --- | --- |
| DTP3 Coverage (%) | -0.016 (0.016) | -0.048, 0.016 |
| Breastfeeding (%) | -0.011 (0.013) | -0.036, 0.014 |
| Health Expenditure (%) | -0.062 (0.065) | -0.190, 0.066 |
| Pneumonia | 0.007 (0.013) | 0.0065, 0.032 |
| Diarrhoea | 0.032 (0.013) | 0.006, 0.057 |

#### Random effects

Table 10 summarizes the posterior estimates of the random effects. The precision estimate for the country-level spatial effect was 77.52, corresponding to a posterior standard deviation of 0.0129, indicating the presence of residual spatial variability in under-five mortality across African countries. The estimated spatial mixing parameter was 0.388 (SD = 0.271), suggesting that both structured and unstructured spatial components contributed to the observed geographical variation. The nonlinear effects of underweight prevalence, stunting, life expectancy, population size, and maternal mortality ratio exhibited relatively high precision values and small posterior standard deviations, reflecting stable nonlinear smooth effects.

**Table 10:** Random effect parameter.

| Random Effect | Precision (Mean) | $\sigma$ (Standard Deviation) |
| --- | --- | --- |
| Gaussian Observation Error | 0.869 | 1.151 |
| Underweight (RW2) | 166.667 | 0.006 |
| Stunting (RW2) | 161.29 | 0.0062 |
| Life Expectancy (RW2) | 163.934 | 0.0061 |
| Population (RW2) | 163.934 | 0.0061 |
| Maternal Mortality Ratio (RW2) | 161.29 | 0.0062 |
| Spatial Effect (Country-Level BYM2) | 77.52 | 0.0129 |
| Spatial Mixing Parameter ($\phi$) | 0.388 | 0.271 |

### Smoothed Spatial Distribution of Under-Five Mortality

Figure 4 displays the posterior mean estimates of under-five mortality obtained from the fitted Gamma Bayesian spatial model. The smoothed estimates still retained the broad geographical pattern observed in the raw mortality map, although local fluctuations were reduced, resulting in a more coherent representation of the underlying spatial risk.

**Figure 4:**
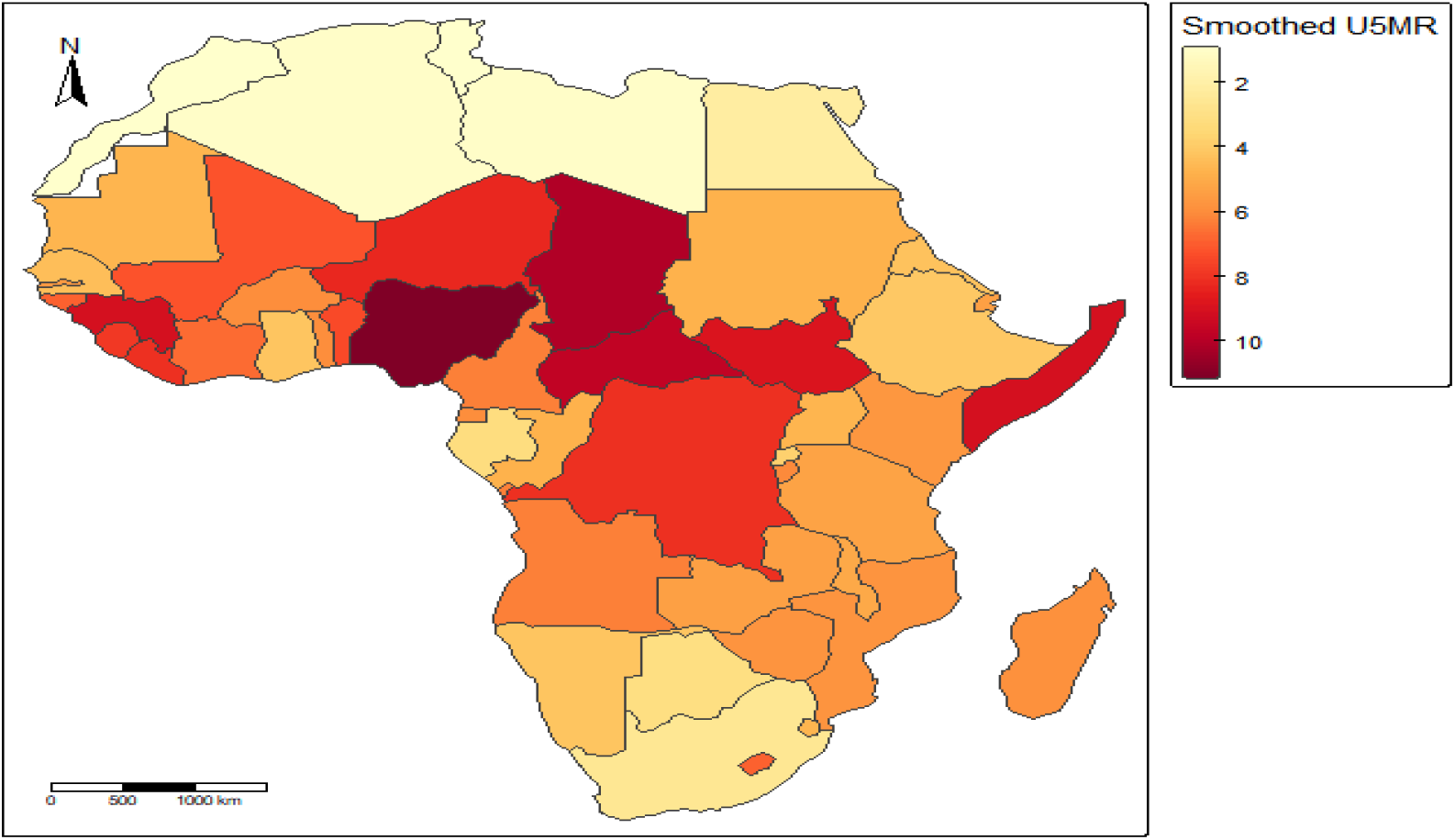
Posterior mean estimates of under-five mortality obtained from the Gamma Bayesian spatial model in Africa, 2024

High mortality levels remained concentrated within West, Central, and parts of East Africa, with Nigeria, Chad, Cameroon, the Democratic Republic of Congo, Somalia, and South Sudan exhibiting the highest posterior mortality estimates. In particular, Nigeria continued to demonstrate the greatest mortality burden, followed by neighbouring countries within the West and Central African subregions. Countries such as Benin, Guinea, Sierra Leone, Liberia, and the Central African Republic also maintained relatively high mortality estimates.

In contrast, countries located in North Africa, including Tunisia, Libya, Morocco, Algeria, and Egypt, consistently exhibited lower posterior mortality estimates. In addition, several countries in Southern Africa displayed comparatively reduced mortality burdens after spatial smoothing.

Compared with the raw prevalence map, the smoothed estimates showed less abrupt variation between adjacent countries, suggesting that the Bayesian model effectively accounted for random fluctuations and borrowing of information from neighbouring areas. Not with standing, the persistence of elevated mortality levels in West, Central, and parts of East Africa indicates that geographical inequalities in child survival remain across the continent despite adjustment for observed risk factors and spatial dependence.

### Residual Spatial Autocorrelation Analysis

Figure 5 displayed the residual spatial autocorrelation analysis based on Moran’s I statistic. The estimated Moran’s I value was 0.074, which was slightly higher than the expected value under spatial randomness (−0.021). However, the corresponding *Z*-score (0.953) and p-value (0.341) which indicate that the residual spatial autocorrelation was not statistically significant. This suggests that the fitted Gamma Bayesian spatial model adequately captured the underlying spatial structure of under-five mortality in Africa, leaving no evidence of residual clustering after accounting for observed covariates, nonlinear effects, and spatial random effects.

**Figure 5:**
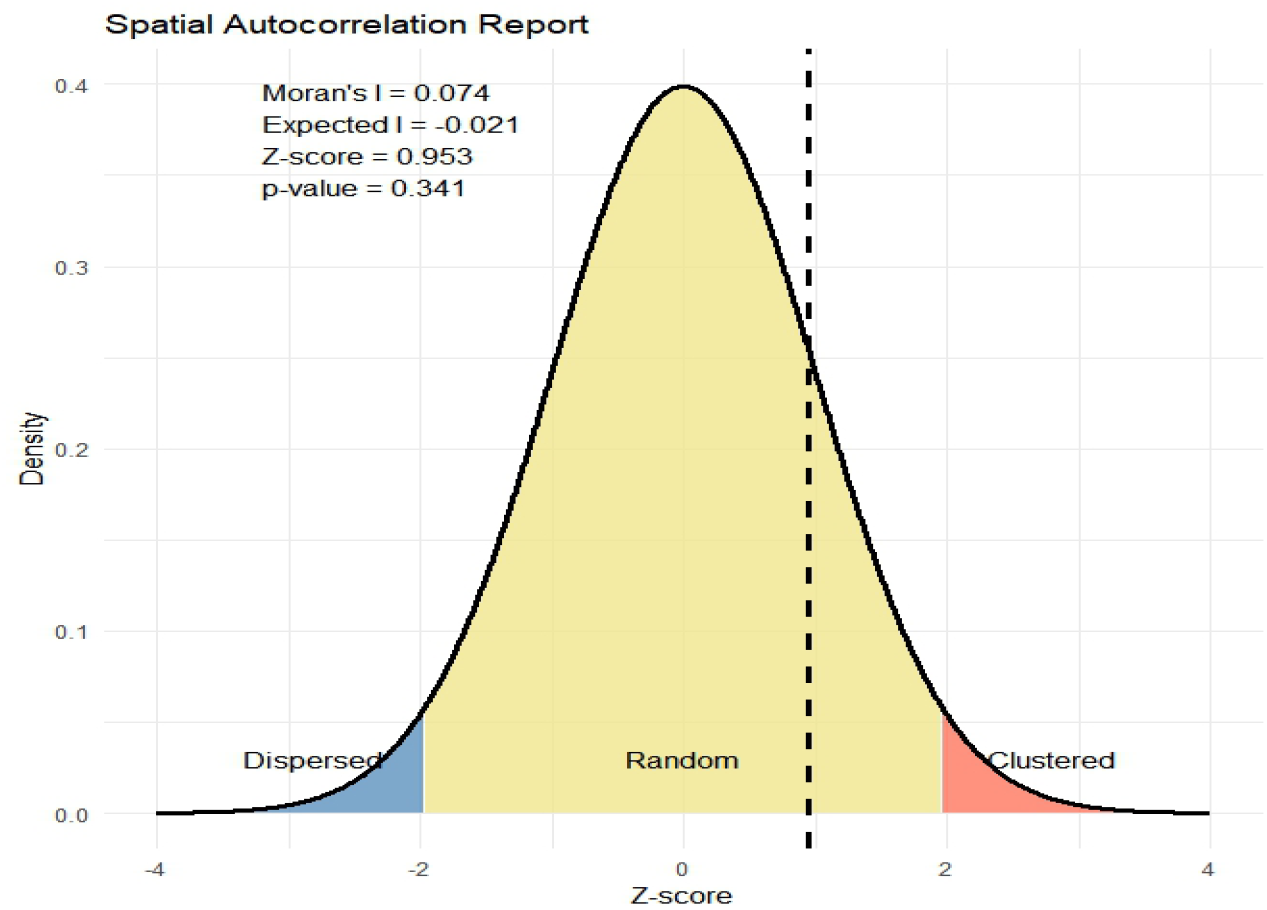
Residual spatial autocorrelation of under-five mortality following Bayesian Gamma spatial modelling.

### SDG 3.2 Attainment Status and Exceedance Risk

Table 11 shows the classification of African countries according to their progress towards achieving SDG 3.2. Out of the 49 countries included in the analysis, only five countries which accounted for almost 10.2% had already achieved the SDG target, while four countries (8.2%) were classified as being near achievement. In contrast, 17 countries (34.7%) were categorized as moderate risk, and nearly half of the continent i.e 23 countries which is almost 47% were identified as high-risk countries, indicating substantial challenges in attaining the SDG target by 2030.

**Table 11:**
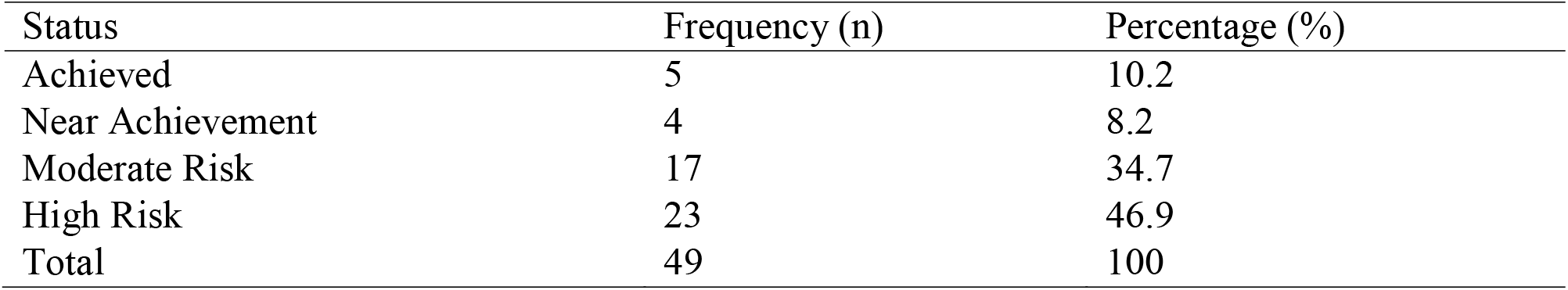
SDG 3.2 Attainment Classification.

| Status | Frequency (n) | Percentage (%) |
| --- | --- | --- |
| Achieved | 5 | 10.2 |
| Near Achievement | 4 | 8.2 |
| Moderate Risk | 17 | 34.7 |
| High Risk | 23 | 46.9 |
| Total | 49 | 100 |

The spatial distribution of SDG attainment categories is presented in Figure 6. Countries that had already achieved the target were predominantly located in North Africa, whereas high-risk countries were concentrated in West, Central, and parts of East Africa. Moderate-risk countries were mainly distributed across Southern and Eastern Africa, while countries approaching attainment were sparsely distributed across the continent. The observed pattern highlights considerable geographical disparities in progress towards SDG 3.2 and implies that the burden of under-five mortality remains disproportionately concentrated in selected regions of Africa.

**Figure 6:**
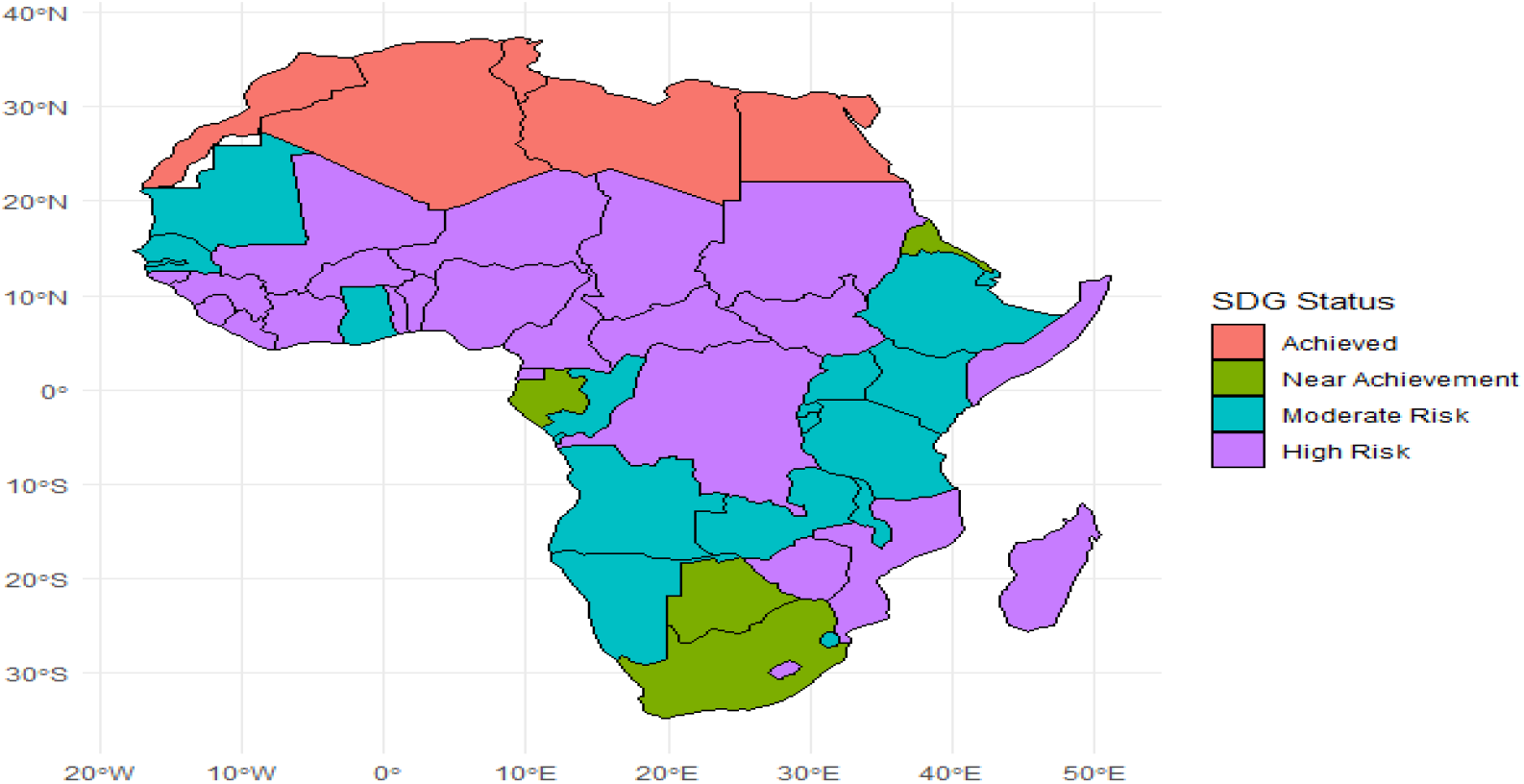
Spatial distribution of SDG 3.2 attainment status for under-five mortality in Africa, 2024.

### SDG Gap Analysis and Country Ranking

Table 12 revealed the ten countries furthest from attaining SDG 3.2. Nigeria exhibited the largest SDG gap (9.1%), followed by Niger (8.6%) and Somalia (7.6%). Chad and South Sudan each recorded a gap of 7.2%, while Guinea, the Democratic Republic of Congo, Sierra Leone, the Central African Republic, and Liberia also remained above the SDG threshold of 2.5%. These findings indicate that progress towards reducing under-five mortality remains particularly challenging in several countries within West, Central, and East Africa.

**Table 12:** Ten worst performing Countries.

| Country | U5MR (%) | SDG Gap(%) |
| --- | --- | --- |
| Nigeria | 11.6 | 9.1 |
| Niger | 11.1 | 8.6 |
| Somalia | 10.1 | 7.6 |
| Chad | 9.7 | 7.2 |
| South Sudan | 9.7 | 7.2 |
| Guinea | 9.2 | 6.7 |
| Democratic Republic of the Congo | 9 | 6.5 |
| Sierra Leone | 9 | 6.5 |
| Central African Republic | 9 | 6.5 |
| Liberia | 8.6 | 6.1 |

**Table 13:**
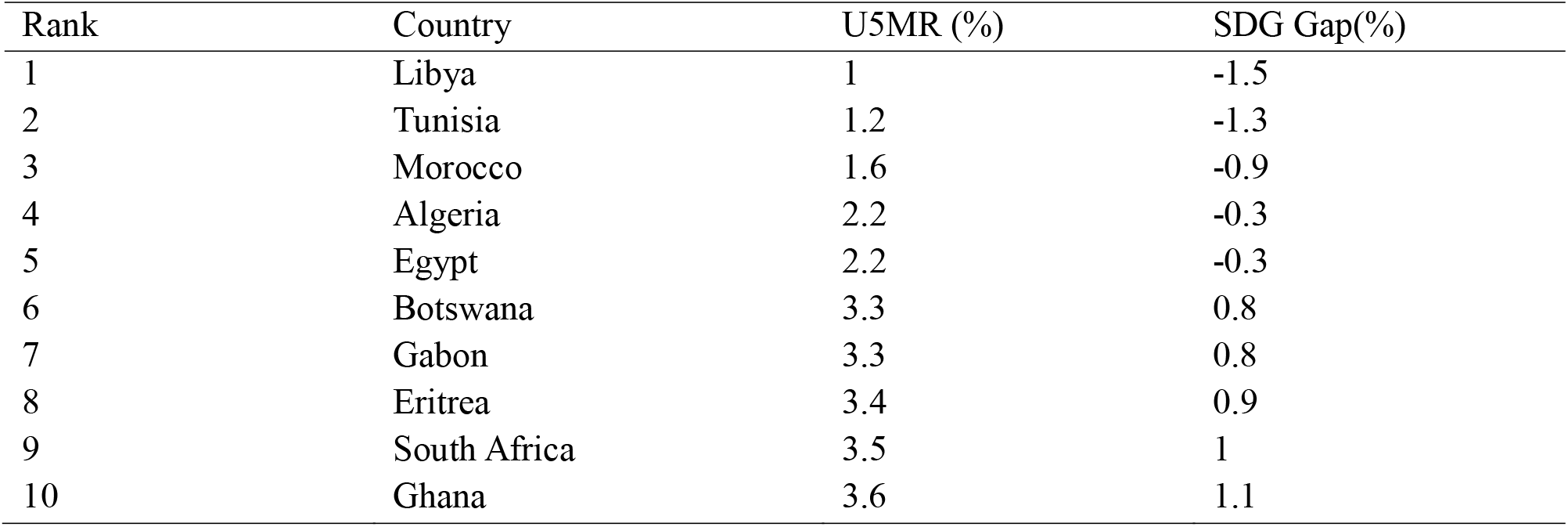
Top 10 Best Performing Countries.

### Continental Inequality in Under-Five Mortality

The result from Table 14 shows the inequality measures for under-five mortality across Africa. The average under-five mortality rate was 5.64%, with a standard deviation of 2.61%. The coefficient of variation was 46.27%, indicating considerable relative variability in mortality levels among African countries. Similarly, the estimated Gini coefficient of 0.263 suggests a moderate degree of inequality in the distribution of under-five mortality across the continent, implying that progress in reducing child mortality has not been uniformly shared among African countries.

**Table 14:** Inequality Indices.

| Indicator | Value |
| --- | --- |
| Mean U5MR (%) | 5.64 |
| Standard Deviation | 2.61 |
| Coefficient of Variation (%) | 46.27 |
| Gini Coefficient | 0.263 |

### Regional Inequalities in Under-Five Mortality

Table 15 show summary regional disparities in under-five mortality across Africa. West Africa recorded the highest average under-five mortality rate (7.05%), followed closely by Central Africa (6.89%), whereas North Africa exhibited the lowest mean mortality level (1.64%) and was the only region with a negative mean SDG gap (−0.86). West and Central Africa also demonstrated the largest average SDG deficits and near-certain probabilities of exceeding the SDG 3.2 threshold. In contrast, North Africa had the lowest exceedance probability (0.082), indicating a substantially lower risk of failing to achieve the target.

**Table 15:** Regional Inequality and exceedance probability.

| Region | Mean U5MR (%) | SD | Median U5MR (%) | Minimum (%) | Maximum (%) | Mean SDG Gap | Exceedance Probability |
| --- | --- | --- | --- | --- | --- | --- | --- |
| West Africa | 7.05 | 2.58 | 7.3 | 3.6 | 11.6 | 4.55 | 1 |
| Central Africa | 6.89 | 2.54 | 6.8 | 3.3 | 9.7 | 4.39 | 0.983 |
| East Africa | 5.5 | 2.24 | 4.8 | 3.4 | 10.1 | 3 | 0.996 |
| Southern Africa | 4.82 | 1.07 | 4.85 | 3.3 | 6.5 | 2.32 | 0.938 |
| North Africa | 1.64 | 0.56 | 1.6 | 1 | 2.2 | -0.86 | 0.082 |

**Figure 7:**
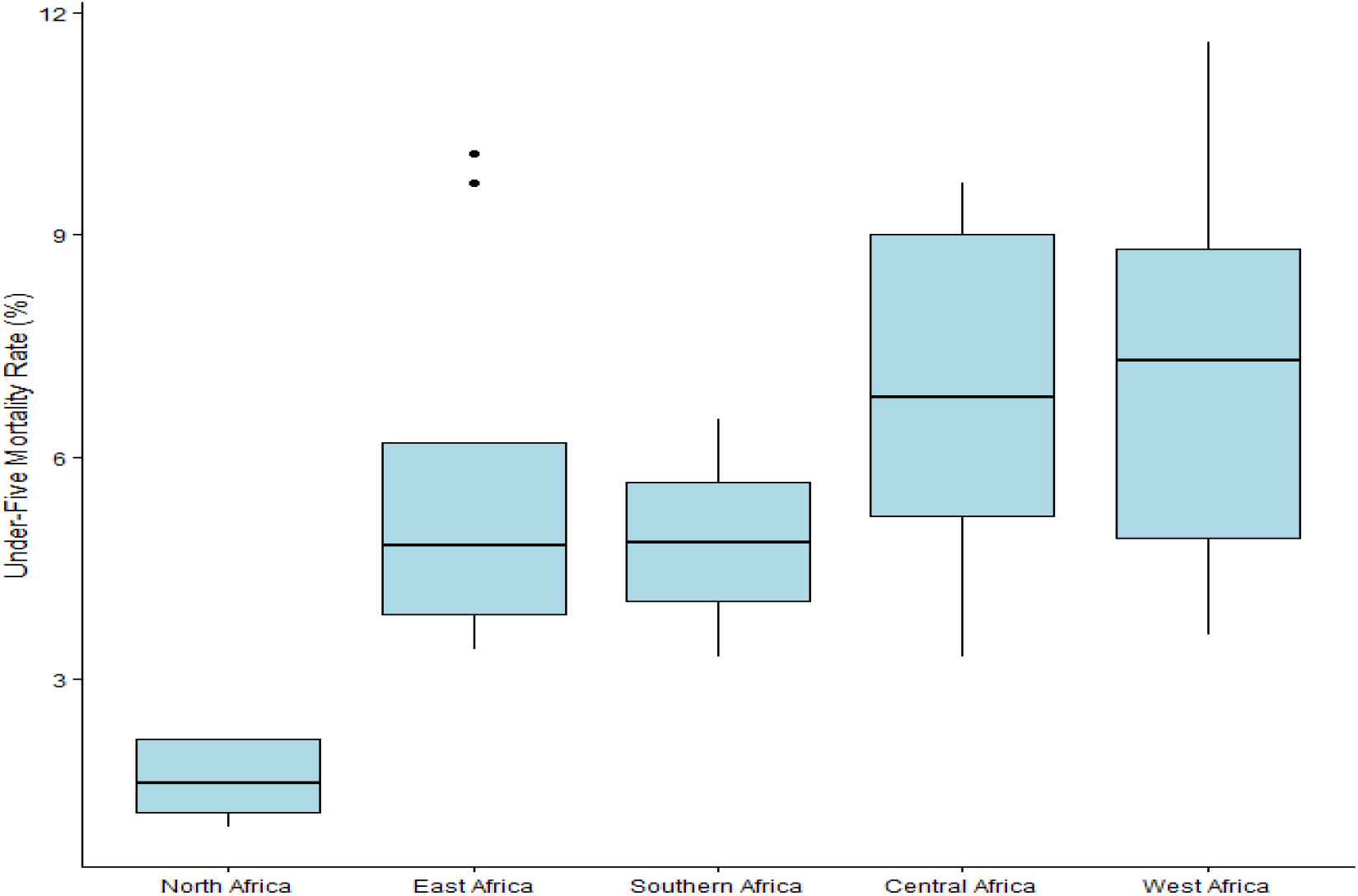
Regional Variation of Under-five Mortality Across Africa

### SDG Failure Index

Figure 8 depicts the spatial distribution of the SDG Failure Index (SFI) across Africa countries. Cleared geographical disparities were observed, with countries in West Africa, some part of East Africa and Central Africa exhibiting the highest priority for intervention. High-priority countries were concentrated in a corridor extending from Nigeria, Niger, Chad, Cameroon, the Democratic Republic of Congo, and Somalia, indicating areas where both the magnitude of the mortality burden and the probability of failing to attain SDG 3.2 remain substantial high.

**Figure 8:**
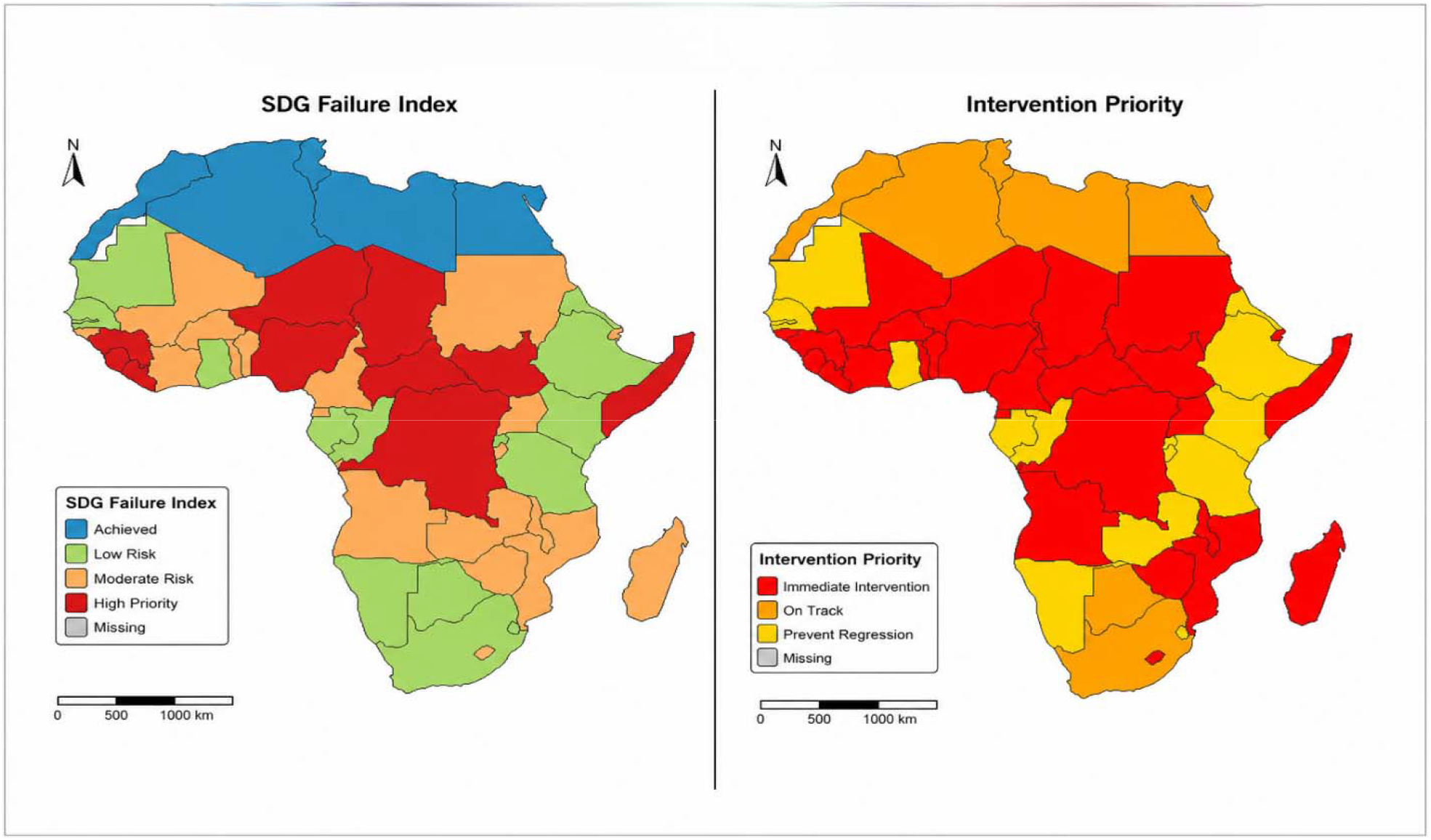
SDG Failure Index and intervention priority Across Africa

In contrast, countries in North Africa, including Libya, Tunisia, Morocco, Algeria, and Egypt, were classified as having already achieved the SDG target and shown the lowest failure indices. Several countries in Southern and Eastern Africa were categorized as low or moderate risk areas, reflecting comparatively better child survival outcomes but continued vulnerability to missing the SDG target.

In overall, the SFI map highlights a distinct north-south divide in progress towards SDG 3.2 and identifies countries where intensified efforts to reduce preventable under-five deaths may yield the greatest gains.

### Priority Intervention Classification and Bayesian Probability of SDG 3.2 Attainment by 2030

Table 16 presents the Priority Intervention Index identified two countries (4.1%) as requiring immediate intervention, while four countries (8.2%) were categorized as moderate priority areas. Most countries (87.8%) were classified as low priority, reflecting either relatively low mortality burdens or sustained reductions in under-five mortality. Taken together, these findings suggest that current rates of decline are insufficient for most African countries to achieve SDG 3.2 by 2030 without accelerated progress.

**Table 16:** Priority Intervention Classification.

| Priority Class | Frequency (n) | Percentage (%) |
| --- | --- | --- |
| High Priority | 2 | 4.1 |
| Moderate Priority | 4 | 8.2 |
| Low Priority | 43 | 87.8 |
| Total | 49 | 100 |

Figure 9 depict country specific probabilities of attaining SDG Target 3.2 by 2030 under current mortality reduction trajectories while accounting for uncertainty in the posterior mortality estimates. Out of 49 countries, only five countries Morocco, Tunisia, Libya, Algeria, and Egypt were classified as likely to achieve the target, with probabilities ranging from 0.85 to 1.00. Three countries, namely Botswana, South Africa, and Gabon, were identified as possible achievers. In contrast, the remaining 41 countries exhibited probabilities below 0.20 and were therefore considered unlikely to attain SDG 3.2 by 2030. Countries with the lowest probabilities were located in West, Central, and some from Eastern Africa, including the Democratic Republic of the Congo, Somalia, Chad, Niger, Nigeria, and Sudan.

**Figure 9.**
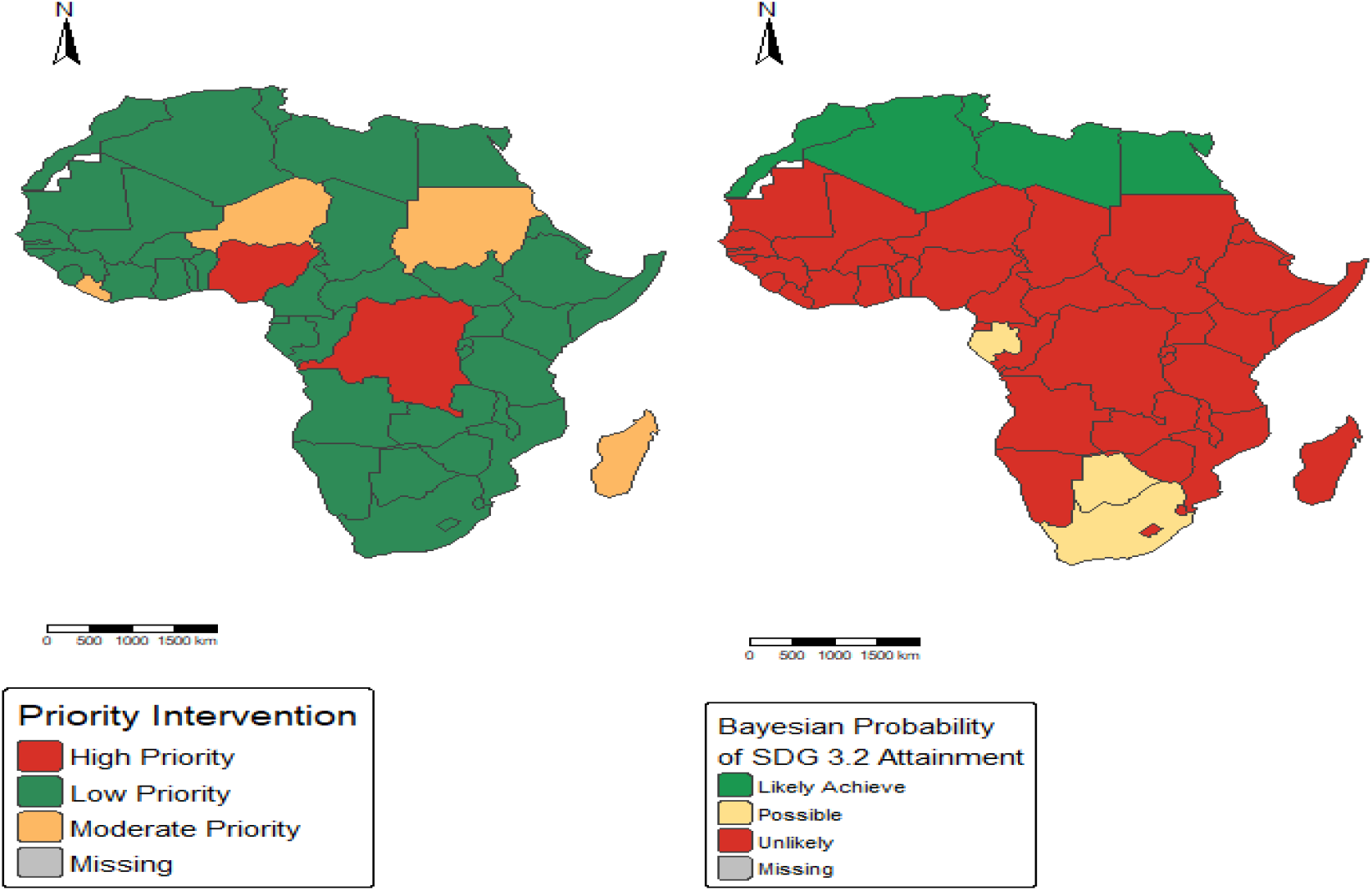
Priority Intervention Classification and Bayesian Probability of Attaining SDG 3.2 by 2030 Across Africa

## Discussion of Findings

This research studied which Africa countries risk missing SDG 3.2 using UNICEF 2024 Data to assess their achievements towards SDG 3.2. The study identified an obvious spatial inequality in under-five mortality across Africa countries and revealed that progress towards Sustainable Development Goal (SDG) 3.2 remains highly uneven across the continent. Unlike previous continental studies that focused primarily on mortality estimation and trend analysis, this study integrated Bayesian exceedance probability mapping, SDG gap assessment, SDG Failure Index construction, and priority intervention classification to identify which countries are most at risk of failing SDG 3.2.

The significant positive Moran’s I statistic observed from the analysis indicates that under-five mortality is not randomly distributed but exhibits clear geographical clustering, with neighbouring countries have tendency of similar under-five mortality burdens. This finding aligns with the continental mapping conducted by Golding et al., 2017, who reported persistent concentrations of high child mortality across West and Central Africa despite considerable reductions in mortality rates between year 2000 and 2015[7]. Golding et al., 2017 also observed that several countries in these regions would require unprecedented rates of mortality decline to achieve SDG child survival targets, this suggest that spatial inequalities in child health outcomes are deeply rooted and difficult to eliminate through general intervention alone.

The spatial pattern observed from the analysis in this study reinforces evidence from the Global Burden of Disease ( Paulson et al., 2021) which showed that although global under-five mortality declined substantially from 71.2 deaths per 1,000 live births in 2000 to 37.1 deaths per 1,000 live births in 2019, progress remained markedly slower in many countries especially countries in sub-Saharan Africa [11]. The GBD study estimated that larger proportion of countries that remain off-track for achieving SDG 3.2 are more in sub-Saharan Africa, which highlight the persistence of regional disparities in child survival. The clustering of high under-five mortality observed in countries such as Nigeria, Niger, Chad, Somalia, South Sudan, Guinea, Sierra Leone, the Democratic Republic of Congo, and the Central African Republic therefore reflect broader continental patterns rather than isolated national challenges.

One of the most important findings of this study is that almost half of African countries remain classified as high-risk countries of under-five mortality based on the SDG attainment framework. This finding aligns closely with recent evidence that many countries remain unlikely to achieve SDG 3.2 by 2030 under existing strategies and trajectories. Using a realist synthesis of global evidence, Qian, (2025) who reported that 59 countries worldwide remain at risk of missing the SDG target for child mortality and that the burden is disproportionately concentrated in sub-Saharan Africa[12]. According to Qian (2025), achieving SDG 3.2 will require stronger governance structures, improved accountability mechanisms, prioritization of effective interventions, and increased investment in community-based health programs. The findings from this study provide empirical support for this argument by identifying specific countries where intervention efforts should be intensified if the SDG 3.2 target is to be achieved.

Another important contribution of this study is the incorporation of Posterior exceedance probability within the SDG monitoring framework. The previous studies have primarily focused on estimating mortality levels and forecasting future trends (Golding et al., 2017; Ebrahimi et al., 2023), this study quantified the posterior probability that each country exceeds the SDG threshold of 25 deaths per 1,000 live births[7,13]. Countries including Nigeria, Niger, Chad, Somalia, and South Sudan exhibited exceedance probabilities approximating to one, indicating an almost certain likelihood of remaining above the SDG target. However, countries such as Tunisia, Libya, Morocco, Algeria, and Egypt demonstrated very low exceedance probabilities. This approach extends existing SDG monitoring methods because it incorporates uncertainty into decision-making which would allow policymakers to distinguish between countries that are merely above the threshold and those with an extremely high probability of failing to meet the target. The priority intervention classification countries requiring urgent policy responses from those where gains should primarily be sustained. Similar risk-based prioritization frameworks have been proposed in global health planning to optimize allocation of scarce resources and maximize population health gains[14]. The SDG Failure Index and Priority Intervention Index proposed in this study were designed to provide an operational framework for identifying countries requiring accelerated progress towards SDG 3.2. Composite indicators have increasingly been advocated for monitoring multidimensional development challenges because they synthesize information from multiple dimensions into interpretable metrics for decision-making [15]. Likewise, recent SDG monitoring frameworks emphasize the importance of identifying countries that are simultaneously experiencing substantial development deficits and insufficient rates of progress[16].

The regional analyses revealed pronounced disparities across Africa sub-regions. West Africa recorded the highest average under-five mortality burden, followed closely by Central Africa, whereas North Africa demonstrated the lowest mortality levels and the smallest posterior exceedance probabilities. These findings closely mirror those reported by Golding et al. (2017), who opined that West and Central Africa as the regions contributing most significantly to the continental burden of under-five child mortality. The decline in mortality and attainment of SDG 3.2 by North African countries may reflect stronger health systems, broader immunization coverage, improved maternal and child health services, greater socioeconomic development, and lower levels of conflict relative to many countries in sub-Saharan Africa. Such differences highlight the importance of considering geographical context when evaluating progress towards SDG 3.2.

The inequality analysis conducted in this study provides further insight into continental disparities. The coefficient of variation of approximately 46% and a Gini coefficient of 0.263 indicate reasonable level of inequality in under-five mortality across African countries. Although reductions in average mortality occurred over recent decades, the persistence of wide inequalities suggests that improvements have not been evenly distributed. This result supports the argument of Griggs et al., (2014) that sustainable development should not be evaluated solely through aggregate progress but through integrated approaches that simultaneously address human wellbeing and equity[17]. Similarly, Cernev & Fenner, (2019) emphasized that SDG 3 represents a foundational goal whose achievement could bring about progress across multiple development areas. Consequently, persistent inequalities in child survival may limit collective sustainable development objectives throughout the continent of Africa[18].

The correlation analyses identified strong positive associations between under-five mortality and maternal mortality, stunting prevalence, and population burden, while life expectancy exhibited a strong inverse relationship with under-five mortality. Similar result was reported by Paulson et al., in 2021 who opined that countries with weaker health systems and poorer healthcare access continue to experience disproportionately high mortality burdens. The strong negative association with life expectancy observed in this study further supports the position of other scholars that under-five mortality serves as a broad indicator of population health and societal wellbeing [11].

Within the Bayesian spatial model, diarrhoea emerged as the only significant covariate with 95% credible interval that excluded zero, indicating a robust association with under-five mortality after accounting for spatial dependence and uncertainty. This finding is consistent with the Paulson et al., (2021) study, who identified diarrhoea diseases among the leading causes of mortality in under-five children worldwide and highlighted their continued importance in low-income settings[11]. Despite progress in expanding access to oral rehydration therapy, vaccination, sanitation, and safe water, diarrhoea diseases remain a significant contributor to preventable child deaths across many African countries. Based on this persistent relationship further mortality reductions will require continued investment in water, sanitation, hygiene infrastructure, and integrated child health programmes.

The methodological findings of this study are also noteworthy. The Gamma Bayesian INLA model demonstrated superior performance compared with alternative model specifications such as Gaussian and exponential, with lowest DIC and WAIC values while effectively eliminating residual spatial autocorrelation. This indicates that the model successfully captured the underlying spatial structure of under-five mortality across Africa and produced reliable estimates of risk. The successful application of Bayesian INLA spatial modelling, exceedance posterior probability mapping, SDG gap analysis, and the SDG Failure Index provide a comprehensive analytical framework for evaluating progress towards SDG 3.2. Unlike conventional descriptive approaches, this framework identifies not only where mortality remains high but also where the risk of SDG failure is greatest.

The probability of attainment analysis showed that only five African countries were likely to achieve SDG 3.2 by 2030 under current trajectories. This finding is broadly consistent with projections reported by the United Nations Inter-agency Group for Child Mortality Estimation, which suggest that many sub-Saharan African countries remain off-track to meet child survival targets without significant reductions in mortality [2]. Introducing posterior probabilities into SDG monitoring provides a more informative measure than point estimates alone because it explicitly accounts for uncertainty surrounding future progress.

Finally, the findings indicate that significant progress has been made in reducing under-five mortality across Africa, consistent with global trends reported by Paulson et al., (2021). However, major geographical inequalities remain, and many countries continue to face considerable challenges in achieving SDG 3.2 by 2030. The concentration of under-five mortality burden within West, Central, and parts of East Africa, combined with high exceedance posterior probabilities and large SDG gaps, suggests that accelerated and geographically targeted interventions will be required. Strengthening health systems, improving access to essential child health services, curtail diarrhoea disease burden, and reducing inequalities in healthcare access should remain central priorities for policymakers seeking to accelerate progress towards ending preventable child deaths across Africa Countries.

## Conclusion

This study revealed significant geographical inequalities in under-five mortality across Africa countries and revealed that progress towards SDG 3.2 remains highly uneven. Significant spatial clustering was observed, with the greatest burden concentrated in West, Central and parts of East Africa. Bayesian exceedance probability mapping showed that many countries continue to face a high likelihood of remaining above the SDG threshold of 25 deaths per 1,000 live births. The newly proposed SDG Failure Index and Priority Intervention Index provided additional insight into the magnitude of U5MR deficits and the urgency of policy responses, while Bayesian posterior predictive simulations suggested that less than one in eight African countries are likely to attain SDG 3.2 by 2030 under current trajectories. These findings underscore the need for geographically targeted interventions, strengthened health systems and sustained investments in child survival programmes if preventable under-five deaths are to be substantially reduced and sustained across the continent.

## Data Availability

All data supporting the findings of this study are available within the paper.
The datasets used in this study are publicly available upon request from the UNICEF.

https://s0mjq.mjt.lu/lnk/AVYAAJb6mDIAAc5R6RIAA56LP3gAAYDOtb8Anb2nACX-xgBp753aTSZ2YKhTSgO3GWEWK8dGuAAayHg/1/EsK8Ftg9LJkpF8OE8eQmNA/aHR0cHM6Ly9kYXRhLnVuaWNlZi5vcmcvcmVzb3VyY2VzL2FmY3NjLTIwMjYvP3V0bV9jYW1wYWlnbj1BZnJpY2ElMjdzJTIwQ2hpbGRyZW4lMjBTdGF0aXN0aWNhbCUyMENvbXBlbmRpdW0lMjAlMjhBZkNTQyUyOSUyMDIwMjYuJnV0bV9tZWRpdW09ZW1haWwmdXRtX3NvdXJjZT1NYWlsamV0

## Strengths and Limitations

This study combines Bayesian spatial disease mapping with SDG monitoring tools to provide a comprehensive assessment of under-five mortality in Africa countries. The introduction of the SDG Failure Index, Priority Intervention Index, and Bayesian probability of SDG attainment by 2030 extends conventional mortality mapping by incorporating uncertainty and recent progress in mortality reduction. In addition, the use of publicly available and comparable datasets permits reproducibility and facilitates future updates as new data become available.

The findings should, however, be interpreted in light of certain limitations. The analysis was undertaken at the national level and may therefore obscure important subnational heterogeneity in child mortality. Furthermore, the Priority Intervention Index was proposed as a pragmatic decision-support metric and its classification thresholds have not been externally validated.

Future studies based on longitudinal and subnational data may help refine the proposed framework and assess its applicability in other settings.

## Funding

The authors received no specific grant from any funding agency in the public, commercial, or not-for-profit sectors for this study.

## Conflict of Interest

The authors declare that they have no competing interests.

## Abbreviations

UNICEF: United Nations Children Education Funds
INLA: Integrated Nested Laplace Approximation
AIC: Akaike Information Criterion
WAIC: Watanabe Akaike Information Criterion
BIC: Bayesian Information Criterion

## Ethics approval and consent to participate

The data is already in public domain therefore Human Ethics and Consent to Participate declarations: not applicable

## Consent for publication

Not Applicable

## Availability of data and materials

All data supporting the findings of this study are available within the paper.

The datasets used in this study are publicly available upon request from the UNICEF.

## Competing Interests

No conflict of interest.

## Authors’ contributions

O.D.M: Conceptualisation, methodology, data curation, formal analysis, software, validation, visualisation, data verification, writing - original draft.

M.K.A: Methodology, supervision, validation, data verification, data interpretation, writing - review & editing.

E.E.E: Literature review, data interpretation, validation, writing - review & editing.

## Acknowledgements

Not Applicable

## Notes

### Competing Interest Statement

The authors have declared no competing interest.

### Author Declarations

The study used UNICEF data only which is openly available human data that were originally located at:https://s0mjq.mjt.lu/lnk/AVYAAJb6mDIAAc5R6RIAA56LP3gAAYDOtb8Anb2nACX-xgBp753aTSZ2YKhTSgO3GWEWK8dGuAAayHg/1/EsK8Ftg9LJkpF8OE8eQmNA/aHR0cHM6Ly9kYXRhLnVuaWNlZi5vcmcvcmVzb3VyY2VzL2FmY3NjLTIwMjYvP3V0bV9jYW1wYWlnbj1BZnJpY2ElMjdzJTIwQ2hpbGRyZW4lMjBTdGF0aXN0aWNhbCUyMENvbXBlbmRpdW0lMjAlMjhBZkNTQyUyOSUyMDIwMjYuJnV0bV9tZWRpdW09ZW1haWwmdXRtX3NvdXJjZT1NYWlsamV0

